## Supplementary material for "Performance of deep-learning based approaches to improve polygenic scores"

**Supplementary Note 1: details of phenotype simulation involving epistasis**

Simulating phenotypes made up from nonlinear genetic effects involves additional parameters and difficulties, relative to phenotypes due to purely additive effects. There are two parameters that need to be considered for phenotypes involving epistatic effects. The first parameter is the number of SNPs that have a causal effect on the phenotype *c*, which would be the only parameter needed for simulating additive phenotypes. However, there is also a second parameter *v*, which controls the number of interactions made up from the *c* SNPs. Preliminary analyses indicated that while *v* had an impact on the overall accuracy, it did not seem to influence the preference between the linear and nonlinear methods, therefore we set *v* = *c* (i.e. all causal SNPs have an interaction).

To reduce the search space, only two phenotypes were simulated, one where the phenotype was due to entirely four-way interactions and another one entirely due to additive effects. Thus, the raw genetic values *GV* for individual *j* were calculated as

$$GV^j = \sum_i^v X_i^j \beta_i \quad ; \quad \beta_i \sim N(0,1),$$

where $\beta_i$ are the SNP (interaction) effects drawn from a standard normal distribution, and $X_i^j$ denotes the *v* randomly selected combinations of SNPs selected to be causal. Thus, $X_i^j$ was defined as

$$X_i^j = \prod_d^{D_i} SNP_{dj},$$

where $SNP_{dj}$ is the genotype count for the *d*th SNP in the *i*th fourth-order interaction.

To simulate phenotypes with an additive genetic architecture with a predefined heritability, the proportion of phenotypic variance due to genetic variance ($h^2$), the final phenotype ($y_{sim}$) is the sum of the genetic (*g*) and noise (*e*) components as

$$y_{sim} = g + e,$$

where both $g$ and $e$ are scaled in proportion to the desired $h^2$. The noise component is drawn from a standard normal distribution, with zero mean and a variance of $1 - h^2$

$$e \sim N(0, \sqrt{1 - h^2}).$$

Thus, the noise contributes all the remaining variance not due to $h^2$. The scaled genetic value ($g$) is in turn defined as

$$g = GV * s,$$

where $s$ is a scaling factor given by

$$s = \sqrt{\frac{h^2}{var(GV)}},$$

where $var(GV)$ denotes the sample variance of the genetic values. The above would generate a simulated phenotype, arising from additive genetic effects, with a pre-specified level of $h^2$. However, to generate phenotypes due to a genetic architecture from nonlinear effects, the above formula was modified by adjusting the scaling factor $s$ to be proportionate to the desired apparent additive effect of SNPs. This was accomplished by fitting a multiple linear regression model, and regressing the individual genetic values ($GV$) on the genotype matrix as

$$GV = \mathbf{X}'\beta' + \epsilon,$$

where $\mathbf{X}'$, $\beta'$ and $\epsilon$ denote the genotype matrix of the individual SNPs involved in interactions, the interaction coefficients and a random noise term, respectively. The fitted values from this model ($\widehat{GV}$), approximate the genetic values due to the apparent main effects of the SNPs involved in interactions. By using this new $\widehat{GV}$ a new scaling factor may be derived by

$$s' = \sqrt{\frac{h^2}{var(\widehat{GV})}}.$$

From this point onward, with the exception of using this new scaling factor $s'$, the rest of the simulation steps were identical to the phenotypes due to purely additive effects.

### Supplementary Tables

| 83 Binary or numeric covariates | 115 Multi-level factor covariates |
|---|---|
| Age at survey (yrs), Sex, Time in education (yrs), Smoking status, Smoking amount: unknown type (pack years), Smoking amount: combined (pack years), Smoking amount: combined (cigarettes/day), Alcohol status, Alcohol amount: combined, Alcohol frequency: combined (days/week), History of diabetes, Anti-diabetic drug status, HRT drug status, Anti-hypertensives drug status, Lipid-lowering unspecified drug status, SBP (mmHg), DBP (mmHg), Height (cm), Weight (kg), BMI (kg/m2), Waist (cm), Hip (cm), Waist/hip ratio, Waist/height ratio, Total cholesterol (mmol/l), HDL-C (mmol/l), Non-HDL-C (mmol/l), LDL-C (mmol/l), Triglycerides (mmol/l), Apolipoprotein A1 (g/l), Apolipoprotein B (g/l), Lp(a) (mg/dl), Haematocrit (%), Haemoglobin (g/l), White cell count (x10^9/l), Forced Expiratory Volume (l/1min), CRP (mg/l), Total protein (g/l), Albumin (g/l), Creatinine (μmol/l), Glucose (mmol/l), HbA1c (%), Eosinophils (%), Urine Creatinine (mmol/l), Urine Potassium (mmol/l), Urine Microalbumin (mg/L), Urine Sodium (mmol/l), Cystatin-c (mg/l), Urine Creatinine upper or lower bound, Urine Potassium upper or lower bound, Urine Microalbumin upper or lower bound, Urine Sodium upper or lower bound, Alkaline Phosphatase (iu/l), Alanine Transaminase (also called GPT) (iu/l), Aspartate Aminotransferase (also called GOT or SGOT) (iu/l ), Basophils (%), Calcium (mmol/l), Direct Bilirubin (μmol/l), Gamma-Glutamyl Transferase (iu/l), Hand grip strength left (kg), Hand grip strength right (kg), Insulin-like Growth Factor 1 (nmol/l), Lymphocytes (%), Monocytes (%), Neutrophils (%), Oestradiol (pmol/l), PEF, Phosphate (mmom/L), Platelet Estimate (10^9/l), Pulse (per min), Red blood cell count (10^12/l), Rheumatoid factor (iu/ml), Sex Hormone-Binding Globulin (nmol/l), Total bilirubin (μmol/l), Testosterone (nmol/l), Urea (mmol/l), Uric acid (μmol/l), Vital capacity (l), Vitamin D (25 dihydroxy-vitamin D) (nmol/L), History of chronic obstructive pulmonary disease (COPD), income: annual amount (cont), | Race, Nationality, Level of education reached, Occupation: job, Smoking status: cigarettes, Smoking status: pipes & cigars, Smoking status: unknown type, Smoking status: combined, Alcohol status: combined, History of CHD, History of MI, History of angina, History of other HD, History of stroke, History of ischaemic stroke, History of haemorrhagic stroke, History of TIA, History of PVD, History of diabetes, History of hypertension, History of cardiovascular surgery, History of coronary revascularisation (surgery), History of CABG (coronary artery bypass graft), History of PTCA (percutaneous transluminal angioplasty), History of vascular surgery, History of neoplasm, Drug status: anti-diabetics, Drug status: HRT, Drug status: anti-hypertensives, Drug status: lipid-lowering unspecified, Family history of CHD - parents, Family history of diabetes - parents, Family history of stroke - parents, Status, All cardiovascular, All cardiovascular (fatal), All cardiovascular (non-fatal), All CHD, All CHD (non-fatal), All CHD (fatal), All CHD plus all cerebrovascular, All CHD plus all cerebrovascular (fatal), All CHD plus all cerebrovascular (non-fatal), Myocardial infarction, Myocardial infarction (fatal), Myocardial infarction (non-fatal), CHD death and non-fatal MI, All cerebrovascular, All cerebrovascular (non-fatal), All cerebrovascular (fatal), Ischaemic stroke, Ischaemic stroke (non-fatal), Ischaemic stroke (fatal), Haemorrhagic stroke, Haemorrhagic stroke (fatal), Haemorrhagic stroke (non-fatal), Subarachnoid haemorrhage, Subarachnoid haemorrhage (fatal), Subarachnoid haemorrhage (non-fatal), Unclassified stroke, Unclassified stroke (fatal), Unclassified stroke (non-fatal), All unknown cause (fatal), All non-cardiovascular (fatal), All tumour (fatal), Digestive related cancer (fatal), Lung cancer (fatal), Genitourinary related cancer (fatal), Breast cancer (fatal), All non-tumour non-cardiovascular (fatal), External (violence/suicide/trauma) (fatal), |

| | |
|---|---|
| income: annual amount (cont) | Infectious/bacterial/parasitic (except hepatitis) (fatal), Mental disorder (fatal), Nervous system disorder (fatal), Liver disease (fatal), Respiratory system disease (fatal), Digestive system disease (except liver) (fatal), Renal disease (fatal), COPD and related conditions (fatal), COVID-19 (fatal), Heart failure (fatal), Status (all-cause mortality), All cardiovascular (all-cause mortality), All CHD (all-cause mortality), All CHD plus all cerebrovascular (all-cause mortality), Myocardial infarction (all-cause mortality), All cerebrovascular (all-cause mortality), Ischaemic stroke (all-cause mortality), Haemorrhagic stroke (all-cause mortality), Subarachnoid haemorrhage (all-cause mortality), Unclassified stroke (all-cause mortality), All unknown cause (all-cause mortality), All non-cardiovascular (all-cause mortality), All tumour (all-cause mortality), Digestive related cancer (all-cause mortality), Lung cancer (all-cause mortality), Genitourinary related cancer (all-cause mortality), Breast cancer (all-cause mortality), All non-tumour non-cardiovascular (all-cause mortality), External (violence/suicide/trauma) (all-cause mortality), Infectious/bacterial/parasitic (except hepatitis) (all-cause mortality), Mental disorder (all-cause mortality), Nervous system disorder (all-cause mortality), Liver disease (all-cause mortality), Respiratory system disease (all-cause mortality), Digestive system disease (except liver) (all-cause mortality), Renal disease (all-cause mortality), COPD and related conditions (all-cause mortality), COVID-19 (all-cause mortality), Heart failure (all-cause mortality), History of heart failure, History of respiratory system disease, Drug status: Birth Control Pill, Drug status: vitamin D |

**S Table 1 |** Full list of covariates used in the real data analyses.

| **Target phenotype** | **Exclusion list** |
|---|---|
| cancer | Breast cancer (all-cause mortality), Digestive related cancer (all-cause mortality), Genitourinary related cancer (all-cause mortality), All tumour (fatal), Digestive related cancer (fatal), Lung cancer (fatal), Genitourinary related cancer (fatal), Breast cancer (fatal), Lung cancer (all-cause mortality) |
| hypertension | All cardiovascular, All cardiovascular (fatal), All cardiovascular (non-fatal), History of other HD, Drug status: lipid-lowering unspecified, Lipid-lowering unspecified drug status, History of hypertension, Drug status: anti-hypertensives, Anti-hypertensives drug status |

| | |
|---|---|
| stroke | All cardiovascular, All cardiovascular (fatal), All cardiovascular (non-fatal), History of other HD, Drug status: lipid-lowering unspecified, Lipid-lowering unspecified drug status, History of stroke, History of ischaemic stroke, History of haemorrhagic stroke, History of TIA, Ischaemic stroke, Ischaemic stroke (non-fatal), Ischaemic stroke (fatal), Haemorrhagic stroke, Haemorrhagic stroke (fatal), Haemorrhagic stroke (non-fatal), Unclassified stroke, Unclassified stroke (fatal), Unclassified stroke (non-fatal), Ischaemic stroke (all-cause mortality), Haemorrhagic stroke (all-cause mortality) |
| cvd | All cardiovascular, All cardiovascular (fatal), All cardiovascular (non-fatal), History of other HD, Drug status: lipid-lowering unspecified, Lipid-lowering unspecified drug status |
| diabetes | History of diabetes, History of diabetes, Drug status: anti-diabetics |
| alcohol | Alcohol status: combined, Alcohol status, Alcohol amount: combined, Alcohol frequency: combined (days/week) |
| smoking | Smoking status: cigarettes, Smoking status: pipes & cigars, Smoking status: unknown type, Smoking status: combined, Smoking status, Smoking amount: cigarettes (pack years), Smoking amount: pipes & cigars (pack years), Smoking amount: unknown type (pack years), Smoking amount: combined (pack years), Smoking amount: combined (cigarettes/day) |
| height | Waist/height ratio, Height (cm), BMI (kg/m2) |
| glucose | Glucose (mmol/l) |
| hba1c | HbA1c (%) |

**S Table 2** | List of excluded covariates for specific target phenotypes.

| trait | EFO | category | Internally curated | ICD10 | ICD9 | field | code | Sex |
|---|---|---|---|---|---|---|---|---|
| Hodgkin's disease | EFO_0000183 | cancer | | C81 | 201 | 20001 | 1052 | all |

| | | | | | | | | |
|---|---|---|---|---|---|---|---|---|
| Breast cancer (female) | EFO_0000305 | cancer | | C50 | | 20001 | 1002 | female |
| Ischemic stroke | EFO_0000712 | Cardio-metabolic | | I63 | | 20002 | 1081 | all |
| Melanomas of skin (diagnosis or history) | EFO_0000756 | cancer | | C43 | 172 | 20001 | 1059 | all |
| Prostate cancer | EFO_0001663 | cancer | | C61 | 185 | 20001 | 1044 | male |
| Thyroid cancer | EFO_0002892 | cancer | | C73 | 193 | 20001 | 1065 | all |
| Cancer of tongue | EFO_0003871 | cancer | | C01, C02 | 141 | 20001 | 1011 | all |
| Basal cell carcinoma | EFO_0004193 | cancer | | C44 | | 20001 | 1061 | all |
| Venous thromboembolism | EFO_0004286 | Cardio-metabolic | | I80 | 451 | 20002 | 1068 | all |
| Malignant neoplasm of testis | EFO_0005088 | cancer | | C62 | 186 | 20001 | 1045 | male |
| Cancer of mouth | EFO_0005570 | cancer | | C04, C06 | 144, 145 | 20001 | 1004 | all |
| Skin cancer | EFO_0009259 | cancer | | C43 | 172 | 20001 | 1059 | all |
| Cancer of larynx | EFO_1000354 | cancer | | C32 | 161 | 20001 | 1006 | all |
| Malignant neoplasm of rectum, rectosig | EFO_1000657 | cancer | | C20, C21 | 154 | 20001 | 1021, 1023 | all |

| | | | | | | | | |
|---|---|---|---|---|---|---|---|---|
| moid junction, and anus | | | | | | | | |
| Colon cancer | EFO_1001950 | cancer | | C18 | 153 | 20001 | 1022 | all |
| Cancer within the respiratory system | MONDO_0000376 | cancer | | C30-C39 | 165 | 20001 | 1084 | all |
| Cancer of bronchus; lung | MONDO_0001407 | cancer | | C34 | 162 | 20001 | 1001 | all |
| Cancer of brain | MONDO_0001657 | cancer | | C71 | 191 | 20001 | 1032 | all |
| Major Depressive Disorder | MONDO_0002009 | behavioural | | F33 | | 20002 | 1286 | all |
| Cancer of eye | MONDO_0002236 | cancer | | C69 | 190 | 20001 | 1030 | all |
| Carcinoma in situ of skin | MONDO_0004641 | cancer | | D04 | 232 | 20001 | 1003 | all |
| Cancer of bladder | MONDO_0004986 | cancer | | C67 | 188 | 20001 | 1035 | all |
| type 2 diabetes | MONDO_0005148 | Cardio-metabolic | | E11 | | 20002 | 1223 | all |
| Colorectal cancer | MONDO_0005575 | cancer | | C18 | 153 | 20001 | 1020 | all |
| Cancer of esophagus | MONDO_0007576 | cancer | | C15 | 150 | 20001 | 1017 | all |
| Height | EFO_0004339 | biometric | ht | | | | | all |

| | | | | | | | | |
|---|---|---|---|---|---|---|---|---|
| Fasting glucose | EFO_0004465 | Cardio-metabolic | glucose1 | | | | | all |
| Glycated haemoglobin levels (HbA1c) | EFO_0004541 | Cardio-metabolic | hba1c | | | | | all |

**S Table 3** | List of all 28 phenotypes and their definitions.

| **nonlinear component** | **formula** |
|---|---|
| any data (**X**) | $\Delta = NN_{nonlinear}(\mathbf{X}) - NN_{linear}(\mathbf{X})$ |
| genuine epistasis | Δ(SNP*PRS) - Δ(SNP) |
| genome-wide GxE | Δ(SNP*PRS, Covs) - Δ(SNP*PRS) - Δ(Covs) |
| per-SNP GxE | Δ(SNP*PRS, Covs) - Δ(SNP*PRS) - Δ(Covs) - Δ($PRS_{indi}$, Covs) |

**S Table 4** | Formulae used to infer the various components of the overall nonlinear effect. *NN* is the neural-network function. **X** refers to any data that may be used as input to the NN. *Δ* is the difference between a nonlinear and linear NN model. *SNP* refers to SNP allele dosage data. *SNP*PRS* refers to SNP allele dosage data premultiplied by per-SNP PRS weights. *Covs* are the environmental covariates. $PRS_{indi}$ refers to individual-level PRS profile scores.

| **EFO** | **linear** | **nonlinear** |
|---|---|---|
| EFO_0000183 | 1.631 | 1.747(s) |
| EFO_0000305 | 1.485 | 0.956 |
| EFO_0000712 | 0.362(s) | 0.421(s) |
| EFO_0000756 | 1.341(s) | 1.056 |
| EFO_0001663 | 0.87 | 0.851 |
| EFO_0002892 | 0.921(s) | 0.515 |
| EFO_0003871 | 2.03(s) | 2.107(s) |
| EFO_0004193 | 1.216(s) | 1.157 |
| EFO_0004286 | 0.595 | 0.945 |

| | | |
|---|---|---|
| EFO_0005088 | 1.083(s) | 0.951 |
| EFO_0005570 | 0.388 | 0.679(s) |
| EFO_0009259 | 1.557(s) | 1.305(s) |
| EFO_1000354 | 0.504 | 1.11 |
| EFO_1001950 | 0.859 | 0.838(s) |
| MONDO_0000376 | 0.663 | 0.774(s) |
| MONDO_0001407 | 0.459(s) | 0.573 |
| MONDO_0001657 | -0.472 | 0.859 |
| MONDO_0002009 | 0.386(s) | 0.23 |
| MONDO_0002236 | 1.669 | 1.716 |
| MONDO_0004641 | 0.61(s) | 0.551 |
| MONDO_0004986 | 1.958(s) | 1.562(s) |
| MONDO_0005148 | 0.484(s) | 0.586 |
| MONDO_0005575 | 0.748(s) | 0.868 |
| MONDO_0007576 | 0.888(s) | 0.932(s) |
| EFO_0004339 | 0.955(s) | 0.96 |
| EFO_0004465 | 0.733(s) | 0.838 |
| EFO_0004541 | 0.954(s) | 0.983 |

**S Table 5** | Per phenotype breakdown of results for the SNP-dosage weighting real data scenario. The **linear** column shows the linear NN and the **nonlinear** column shows the nonlinear NN model performance on the test set. *(s)* indicates if the performance evaluated originated from the ‘small’ model ([24, 12, 6] neurons) or the ’large’ model ([100, 50, 25] neurons), a choice made based on the validation set performance. Units are expressed as a fraction relative to the additive baseline, which was the PGS Catalog PGS.

| **pheno** | **linear** | **nonlinear** |
|---|---|---|
| EFO_0000305 | 1.006 | 1.047 |
| EFO_0000712 | 0.06(s) | 0.06 |
| EFO_0000756 | 1.226(s) | 1.242 |
| EFO_0001663 | 0.908 | 0.863 |
| EFO_0002892 | 2.13 | 2.097 |
| EFO_0004193 | 0.952(s) | 0.951 |

| | | |
|---|---|---|
| EFO_0004286 | 0.869 | -0.099 |
| EFO_0009259 | 0.301 | 1.257 |
| EFO_1001950 | 0.943(s) | 0.935(s) |
| MONDO_0000376 | 0.98(s) | 0.998 |
| MONDO_0001407 | -0.211(s) | -0.339(s) |
| MONDO_0001657 | 3.416 | 3.845 |
| MONDO_0002009 | 0.914 | 0.949 |
| MONDO_0004641 | -0.066(s) | 0.705 |
| MONDO_0004986 | -0.285 | -0.369 |
| MONDO_0005148 | 0.982 | 0.985 |
| MONDO_0005575 | 1.216 | 1.222 |
| MONDO_0007576 | 0.703 | 0.703(s) |
| EFO_0004339 | 0.971(s) | 0.993 |
| EFO_0004465 | 0.349(s) | 0.203 |
| EFO_0004541 | 0.619(s) | 0.533 |

**S Table 6** | Per phenotype breakdown of results for the SNP+Covariate real data scenario. The **linear** column shows the linear NN and the **nonlinear** column shows the nonlinear NN model performance on the test set. *(s)* indicates if the performance evaluated originated from the ‘small’ model ([24, 12, 6] neurons) or the ’large’ model ([100, 50, 25] neurons), a choice made based on the validation set performance. Units are expressed as a fraction relative to the additive baseline, which was a multiple regression model that included the same covariates plus the PGS Catalog PGS.

| | **Typical NN task** | **Phenotype prediction** |
|---|---|---|
| **Task challenge** | problem complexity | signal recovery |
| **Sample size** | large | small |
| **noise** | low | high |
| **spatially structured data** | yes | no |

**S Table 7** | Summary of differences between typical tasks where NNs excel at and phenotype prediction.
